# Race-ethnicity and Perceptional Determinants of COVID-19 Vaccination Intentions: A Cross-sectional Study Among Health Workers and the General Population in the San Francisco Bay Area

**DOI:** 10.1101/2021.04.26.21255893

**Authors:** Yingjie Weng, Di Lu, Jenna Bollyky, Vivek Jain, Manisha Desai, Christina Lindan, Derek Boothroyd, Timothy Judson, Sarah B. Doernberg, Marisa Holubar, Hannah Sample, Beatrice Huang, Yvonne Maldonado, George W. Rutherford, Kevin Grumbach, for the California Pandemic Consortium

## Abstract

**Objective:** To compare intention to receive COVID-19 vaccination by race-ethnicity, to identify perceptional factors that may mediate the association between race-ethnicity and intention to receive the vaccine, and to identify the demographic and perceptional factors most strongly predictive of intention to receive a vaccine.

**Design:** Cross-sectional survey conducted from November, 2020 to January, 2021, nested within two longitudinal cohort studies of prevalence and incidence of SARS CoV-2 among the general population and healthcare workers.

**Study Cohort:** 3,161 participants in the Track COVID cohort (a population-based sample of adults) and 1,803 participants in the CHART Study cohort (a cohort of employees at three large medical centers).

**Results:** Rates of high vaccine willingness were significantly lower among Black (45.3%), Latinx (62.5%), Asian (65%), multi-racial (67.2%), and other race (61.0%) respondents than among white respondents (77.6%). Black, Latinx, and Asian respondents were significantly more likely than white respondents to endorse reasons to not get vaccinated, especially lack of trust. Participants’ motivations and concerns about COVID-19 vaccination only partially explained racial-ethnic differences in vaccination willingness. Being a health worker in the CHART cohort and concern about a rushed government vaccine approval process were the two most important factors predicting vaccination intention.

**Conclusions:** Special efforts are required to reach historically marginalized racial-ethnic communities to support informed decision-making about COVID-19 vaccination. These campaigns must acknowledge the history of racism in biomedical research and health care delivery that has degraded the trustworthiness of health and medical science institutions among non-white population and may continue to undermine confidence in COVID-19 vaccines.

---

A successful COVID-19 mass vaccination program requires not only sufficient supply of a safe and effective vaccine and well-organized distribution, but also a willingness of people to get vaccinated. Vaccination campaigns are more effective and equitable if health officials have a robust understanding of vaccine beliefs and motivations, including variation among sub-populations^1^.

National surveys in the US have found that Black and Latinx individuals have more reservations than their white counterparts about COVID-19 vaccination.^2-6^ Lower rates of vaccination among Black and Latinx populations have been attributed to a combination of barriers to access and reduced enthusiasm for vaccination. ^7,8^ We recently reported our findings from a survey of the general population and medical center workers in the San Francisco Bay Area, documenting that in both groups, Black, Latinx, Asian, multi-race and other race respondents had significantly lower intention to get a COVID-19 vaccine than white respondents.^9^ Here, we expand our analyses, comparing peceptional factors including motivations, concerns and worries about COVID-19 vaccination across racial-ethnic groups and the extent to which these factors mediated differences across racial-ethnic groups in intention to get vaccinated. We also present data on perceptational factors and sociodemographic characteristics that were most important in predicting COVID-19 vaccination intention.

## Methods

### Study design and population

We conducted a cross-sectional survey^9^ from late November, 2020 to January, 2021, nested within two longitudinal cohort studies of prevalence and incidence of SARS CoV-2 among (1) community-residing adults (the TrackCOVID study) and (2) medical center employees (the COVID-19 Healthcare Worker Antibody and Reverse Transcription (RT) PCR Tracking study, CHART). The longitudinal cohort studies consisted of volunteer participants receiving biweekly (CHART) to monthly (TrackCOVID) RT-PCR and antibody tests for SARS-CoV-2. The TrackCOVID study recruited randomly selected community members residing in six San Francisco Bay Area counties, with enrollment occurring between July and December, 2020. The CHART Study recruited healthcare and other staff at three medical centers in the San Francisco Bay Area (Stanford Medical Center, Univeristy of California, San Francisco (UCSF) Health, and Zuckerberg San Francisco General Hospital), enrolled from July through November, 2020.

TrackCOVID used an address-based stratified random sampling strategy to select households eligible for study recruitment. Two strata were considered in the sampling scheme to increase statistical efficiency: estimated COVID-19 cases per census tract determined by modeling, and county. Risk of a household was estimated by modeling prevalent cases within census tracts as reported by counties as a function of sociodemographic, occupational, health and poverty characteristics using data from the 2018 American Community Survey and UCSF Health Atlas.^10^ One adult from each randomly selected household was eligible for participation. The CHART study recruited adults employed in diverse occupations working at the three medical centers.

Participants in both cohorts were sent an electronic survey about COVID-19 vaccination with Research Electronic Data Capture Software (REDCap). In the TrackCOVID cohort, surveys were provided in Spanish and Chinese languages for respondents with limited English proficiency. Those who did not respond to the electronic instrument were invited to complete the survey in person during a regular testing visit for the parent study. The survey was administered to TrackCOVID participants from December 14, 2020 to January 15, 2021, and CHART Study participants from November 27 to December 27, 2020. This time frame coincided with the announcement of emergency use authorizations of the Pfizer BNT162b2 (December 11, 2020)^11^ and the Moderna mRNA-1283 (December 18, 2020)^12^ vaccines.

The TrackCOVID study was designated as a public health surveillance study and not human subjects research under 45 CFR 46.102(l) by Stanford University School of Medicine Administrative Panel on Human Subjects in Medical Research and UCSF Institutional Review Board; the CHART study protocol was approved by the UCSF Institutional Review Board and the Stanford University School of Medicine Panel on Human Subjects in Medical Research.

### Survey instruments

Survey items about vaccination were adapted from the NIH Community Engagement Alliance (CEAL) Against COVID-19 Disparities Draft Common Survey^13^ and informed by conceptual models of vaccine hesitancy.^14,15^ Questions were asked about perception of risk of becoming infected with SARS CoV-2, confidence in the vaccine, and motivation to obtain the vaccine, based on the conceptual model of vaccine behavior developed by Brewer et al.^16^

#### Primary Outcome

Our primary outcome was a participant’s high willingness to receive the vaccine. This binary variable was derived from two survey items. The first item asked, “How likely are you to get an approved COVID-19 vaccine when it becomes available?”, using a 1-7 Likert scale with 1 indicating “not at all likely” and 7 “very likely.” Respondents who scored 3 or greater were asked a second question, “How early would you ideally like to receive the COVID-19 vaccine?”, with response options of “I’d like to be among the earliest,” “I’d like to receive it early, but not in the first round of people,” “I’d like to receive it later in the distribution process,” “or “I’d like to wait at least two months to see what the experience is.” Respondents who selected 3 or greater on the first item and answered “I’d like to be among the earliest” or “I’d like to receive it early…” to the second item were categorized as having “high” vaccination willingness.

#### Motivators, Concerns and Worries (Perceptional Factors)

The “motivators” set of items listed reasons for people to want to get a COVID-19 vaccine, asking respondents to rate the importance of each reason on a scale ranging from “Not a reason for me to get the vaccine” to “Most important reason to get the vaccine.” Respondents could pick more than one reason as most important. Responses were then dichotomized to indicate whether that reason was classified as a most important reason. The “concerns” set of items asked about reasons to not get the vaccine, with response options ranging from “Not a reason” to “Most important reason to not get the vaccine.” Responses were dichotomized to indicate whether the concern was a most or moderately important consideration in not getting the vaccine. Two additional items measured degree of worry that a COVID-19 vaccine “Might not stop you from getting COVID” and “Might give you COVID and make you sick,” with responses dichotomized (very or somewhat worried vs. neutral or not at all) (Appendix Table 2). We considered the above perceptional factors as potential mediators for the association between race-ethnicity and vaccine willingness.

#### Sociodemographic Variables

Participants completed a survey at their baseline enrolment visit in the parent cohort study that included questions about socio-demographics. Participants self-identified their race-ethnicity using Office of Management and Budget (OMB) categories, with one item asking about Hispanic/Latinx identity and one item asking about race with response options of White, Black or African American, American Indian or Alaska Native, Asian, Native Hawaiian or Other Pacific Islander, and Other race. Participants could select more than one race. The ethnicity and race items were then combined to create a single variable with mutually exclusive categories (white, Black, Latinx, Asian, mixed race-ethnicity, and other race-ethnicity). Because of the small number of American Indian and Native Hawaiian repondents, these respondents were included in the Other race-ethnicity category. The few resondents who identified as both Hispanic/Latinx and Black were categorized as Black. Other sociodemographic variables included age, gender, occupation, and highest level of education attained.

### Statistical Methods

#### Descriptive analysis

Frequencies and proportions of the social-demographic variables were described for our study cohort. Logistic regression was employed to characterize the association between race-ethnicity and each perceptional factor (motivator, concern, worries), wth calculation of adjusted odds ratios (aOR) and 95% confidence intervals (CI). Regression models adjusted for age, gender, education, and cohort.

#### Mediation analysis

We evaluated whether perceptional factors (motivators, concerns and worries) mediated the association between race-ethnicity and vaccine willingness using Poisson regression. Prevalence ratios (PR) and the corresponding 95% confidence intervals (CI) of vaccine willingness were estimated from the models. Specifically, a step wise Poisson model was performed that first only included cohort enrolled and demographic characteristics as predictors of high vaccine willingness and then, in a second step, added all the perceptional factors including motivators, concerns and worries as predictors. We compared the PRs for race-ethnicity from the two models to identify if perceptional factors mediated the observed race-ethnicity disparity. Complete case analysis was applied for this analysis.

#### Least Absolute Shrinkage and Selection Operator (LASSO) model

Finally, we employed a statistical learning method using LASSO^17^ to identify which motivators, concerns, and sociodemographic features were most important in predicting high vaccine willingness. We first randomly split our sample into training and testing data sets in a 1:1 ratio. We then applied LASSO approaches with 5-fold cross-validations in the training sample and evaluated the performance in the testing sample. Area under curve (AUC) and C-index were calculated as the performance index for both training and testing sets. We evaluated a range of values for the regularization parameter (λ) in the training sample and chose the optimal λ that minimized the change of the performance index to build our final model. Subgroups within a single categorical variable were grouped together using grouped LASSO techniques so that each of the input variables would be either chosen or not chosen as a whole instead of by each individual subgroup within that variable.^18^ For example, we would either select or not select race-ethnicity using this approach, rather than having an indicator for ‘Black’ selected but an indicator of ‘Latinx’ not selected as the predictors of vaccine intention. AUC and its corresponding 95% confidence interval calculated through bootstrapping were reported from the final validation model using the test set. Variable importance were estimated using the absolute value of the β coefficients from the LASSO model in the training set. We ranked the variable importance for each of the selected predictors in a descending order and presented it using a funnel plot. Multiple imputations were performed prior to LASSO regressions to handle variables with missing data.

#### Sensitivity analysis

We performed sensitivity analyses using alternative measures of willingness as the outcome variable. One alternative measure of high willingness was a single item asking respondents to rate their degree of agreement with the statement, “I plan to wait and see how it goes with people who first get the vaccine before getting a vaccine myself.” Respondents who strongly disagreed or disagreed were categorized as having high vaccination willingness. We also created a high hesitancy measure, based on reporting a low likelihood of getting vaccinated.

All statistical analyses was performed using R Statistical Programming Languages, version 4.0.3 and SAS, version 9.4 (SAS Institute, Cary NC).

## Results

All 3,935 participants in the Track COVID study and 2,501 particpants in the CHART study were sent the survey, of whom 3,161 (80.3%) and 1,803 (72.1%) responded, respectively. In both cohorts, those who responded were more likely to be older, white, and highly educated compared to non-responders (Appendix Table 3). Among survey respondents, 120 (2.4%) did not report race-ethnicity and were excluded from the analysis. The total sample used for analysis was 4,964.

Table 1 shows the demographic characteristics of the study sample. The mean age of survey respondents was 47.6 years old (SD: 14.8), 61.4% were female, 58.8% were white and 7.2% did not have a college degree. Among Track COVID respondents, 258 (8.2%) were employed in the healthcare sector.

**Table 1:** Characteristics of survey respondents.

|  | Survey Respondents<br>(N=4964) |
| --- | --- |
|  | n (%) |
| **Age, years** |  |
| 18-39 | 1783 (35.9) |
| 40-64 | 2385 (48.0) |
| >=65 | 787 (15.9) |
| Unknown | 9 (0.2) |
| *Age, years (mean (SD))* | 47.6 (14.8) |
| **Gender** |  |
| Female | 3050 (61.4) |
| Male | 1774 (35.7) |
| Other<sup>1</sup> | 35 (0.7) |
| Unknown | 105 (2.1) |
| **Race-Ethnicity Group** |  |
| White | 2917 (58.8) |
| Black | 139 (2.8) |
| Hispanic/Latinx | 466 (9.4) |
| Asian | 940 (18.9) |
| Multiple races | 259 (5.2) |
| Other | 123 (2.5) |
| Unknown | 120 (2.4) |
| **Education** |  |
| Less than college | 358 (7.2) |
| College<sup>2</sup> | 2195 (44.2) |
| Higher than college | 2240 (45.1) |
| Unknown | 171 (3.4) |
| **Occupation, *Medical Center Workers (CHART study)*** | 1803 (36.3) |
| Physician, advanced practitioner, nurse | 1382 (76.7) |
| Pharmacist, therapist, technician | 217 (12.0) |
| Other medical center occupation | 204 (11.3) |
| **Occupation, *General Population (TrackCOVID study)*** | 3161 (63.7) |
| Employed in health sector | 258 (8.2) |
| Not employed in health sector | 2903 (91.8) |

1. Self-reported gender group: other gender group included trans male, trans female, or genderqueer/gender non-binary
2. Included associate degree (junior college) and bachelor's degree

### Vaccine Willingness

CHART Study participants were more likely than TrackCOVID participants to be highly willing to be vaccinated (83.6% compared with 65.5%). Black, Latinx, Asian, Multi-racial, and Other race respondents were significantly less likely than white respondents in both cohorts to have high willingness to be vaccinated (Table 2).

**Table 2:** High COVID-19 Vaccination Willingness, According to Race-Ethnicity and Study Cohort.

|  | **CHART** |  | **TrackCOVID** |  |
| --- | --- | --- | --- | --- |
|  | Number of Respondents | Percentage Reporting High Vaccine Willingness (95% CI ) | Number of Respondents | Percentage Reporting High Vaccine Willingness (95% CI ) |
| All Respondents | 1803 | 84% (82%-85%) | 3161 | 66% (64%-67%) |
| White | 989 | 89% (87%-91%) | 1927 | 72% (70%-74%) |
| Black/African American | 23 | 65% (46%-85%) | 116 | 41% (32%-50%) |
| Latinx/Hispanic | 154 | 78% (71%-84%) | 312 | 55% (49%-60%) |
| Asian | 365 | 75% (71%-80%) | 575 | 58% (54%-62%) |
| Other Race | 50 | 66% (53%-79%) | 73 | 58% (46%-69%) |
| Multiple Races | 105 | 79% (71%-87%) | 154 | 59% (51%-67%) |

### Association between Sociodemographic and Percepational Factors and Vaccine Willingness

Despite differences in willingness to get vaccinated, reasons to get vaccinated were similar across racial-ethnic groups (Table 3). Latinx and Asian respondents were, however, significantly more likely than white respondents to report “my doctor’s recommendation” as a most important reason.

**Table 3:** Comparison of vaccine motivators and concerns, by race-ethnicity (reference group: white)

| *Outcomes* | *White*<br>*(n=2917)* | *Black*<br>*(n=139)* |  | *Asian*<br>*(n=940)* |  | *Hispanic/Latinx*<br>*(n=466)* |  | *Multiple Races*<br>*(n=259)* |  | *Other Races*<br>*(n=123)* |  |
| --- | --- | --- | --- | --- | --- | --- | --- | --- | --- | --- | --- |
|  | *n (%)* | *n (%)* | *aOR*<br>*[95% CI]* | *n (%)* | *aOR*<br>*[95% CI]* | *n (%)* | *aOR*<br>*[95% CI]* | *n (%)* | *aOR*<br>*[95% CI]* | *n (%)* | *aOR*<br>*[95% CI]* |
| **Reasons to get vaccinated (motivators), % indicating most important reason** |  |  |  |  |  |  |  |  |  |  |  |
| a. I want to protect myself from getting COVID | 2071<br>(71.3) | 102<br>(73.4) | 1.02<br>[0.69, 1.52] | 665<br>(71.0) | 1.18<br>[1.00, 1.40] | 313<br>(67.8) | 0.99<br>[0.80, 1.24] | 168<br>(64.9) | 0.94<br>[0.71, 1.23] | 84<br>(68.3) | 0.98<br>[0.66, 1.47] |
| b. I want to protect my family from getting COVID | 2397<br>(82.7) | 108<br>(78.3) | 0.79<br>[0.52, 1.20] | 780<br>(83.2) | 1.01<br>[0.83, 1.24] | 392<br>(85.0) | 1.19<br>[0.89, 1.58] | 220<br>(84.9) | 1.12<br>[0.78, 1.62] | 97<br>(79.5) | 0.79<br>[0.49, 1.25] |
| c. I want to protect my community from getting COVID | 2044<br>(70.8) | 88<br>(64.2) | 0.73<br>[0.51, 1.05] | 631<br>(67.6) | 0.89<br>[0.76, 1.05] | 323<br>(70.1) | 0.99<br>[0.79, 1.24] | 184<br>(71.0) | 1.00<br>[0.75, 1.33] | 81<br>(66.9) | 0.82<br>[0.55, 1.21] |
| d. I dont believe life will go back to normal until most people get the vaccine | 1837<br>(63.7) | 79<br>(57.2) | 0.75<br>[0.52, 1.06] | 554<br>(59.1) | 0.90<br>[0.77, 1.06] | 275<br>(59.9) | 0.93<br>[0.75, 1.15] | 147<br>(57.0) | 0.84<br>[0.65, 1.10] | 72<br>(59.5) | 0.93<br>[0.63, 1.36] |
| e. I will get the COVID vaccine if my doctor recommends that I get it | 832<br>(28.8) | 52<br>(38.0) | 1.32<br>[0.91, 1.91] | 323<br>(34.5) | 1.52<br>[1.29, 1.79] | 158<br>(34.7) | 1.34<br>[1.08, 1.68] | 74<br>(28.7) | 1.16<br>[0.87, 1.55] | 32<br>(26.2) | 0.95<br>[0.62, 1.44] |
| f. My employer or school will require or expect me to get a vaccine | 551<br>(19.2) | 22<br>(16.4) | 0.83<br>[0.52, 1.34] | 250<br>(27.0) | 1.53<br>[1.28, 1.82] | 105<br>(23.1) | 1.13<br>[0.88, 1.45] | 63<br>(24.6) | 1.25<br>[0.92, 1.71] | 26<br>(21.5) | 1.02<br>[0.64, 1.61] |
| g. My family will want me to get a vaccine | 955<br>(33.2) | 42<br>(31.3) | 0.78<br>[0.53, 1.16] | 324<br>(34.8) | 1.27<br>[1.08, 1.50] | 156<br>(34.2) | 1.10<br>[0.88, 1.38] | 81<br>(31.6) | 1.12<br>[0.84, 1.48] | 41<br>(33.6) | 1.00<br>[0.67, 1.50] |
| **Reasons to not get vaccinated (concerns), % indicating most important or moderately important reason** |  |  |  |  |  |  |  |  |  |  |  |
| a. The vaccine may not stop me from getting COVID | 333<br>(11.5) | 35<br>(25.9) | 2.39<br>[1.58, 3.61] | 180<br>(19.4) | 1.85<br>[1.51, 2.27] | 105<br>(22.9) | 2.04<br>[1.58, 2.64] | 38<br>(14.8) | 1.25<br>[0.86, 1.81] | 24<br>(20.3) | 1.77<br>[1.10, 2.84] |
| b. I might have a bad reaction to the vaccine | 735<br>(25.5) | 62<br>(46.6) | 2.25<br>[1.57, 3.22] | 408<br>(44.0) | 2.23<br>[1.90, 2.61] | 191<br>(41.8) | 1.83<br>[1.48, 2.27] | 93<br>(36.0) | 1.46<br>[1.11, 1.92] | 59<br>(48.4) | 2.49<br>[1.71, 3.61] |
| c. I do not get vaccines in general | 95<br>(3.3) | 18<br>(13.4) | 3.63<br>[2.10, 6.27] | 57<br>(6.2) | 2.03<br>[1.44, 2.88] | 41<br>(9.0) | 2.61<br>[1.75, 3.89] | 15<br>(5.8) | 1.71<br>[0.95, 3.08] | 8 (6.7) | 2.14<br>[1.01, 4.56] |
| d. I do not think I will get COVID, even without getting a vaccine | 50<br>(1.7) | 16<br>(12.0) | 6.86<br>[3.74, 12.6] | 51<br>(5.5) | 3.48<br>[2.31, 5.23] | 25<br>(5.5) | 3.12<br>[1.87, 5.22] | 7<br>(2.7) | 1.41<br>[0.59, 3.37] | 6 (5.1) | 2.59<br>[1.00, 6.66] |
| e. The COVID-19 outbreak is not as serious as some people say it is | 46<br>(1.6) | 14<br>(10.4) | 5.20<br>[2.70, 10.0] | 37<br>(4.0) | 2.52<br>[1.60, 3.96] | 25<br>(5.5) | 3.17<br>[1.88, 5.33] | 7<br>(2.7) | 1.51<br>[0.63, 3.62] | 5 (4.2) | 2.70<br>[1.04, 6.98] |
| f. I do not trust the companies making COVID-19 vaccines | 233<br>(8.1) | 32<br>(23.5) | 3.08<br>[2.00, 4.73] | 103<br>(11.1) | 1.34<br>[1.04, 1.72] | 74<br>(16.2) | 1.85<br>[1.38, 2.48] | 36<br>(14.0) | 1.46<br>[0.98, 2.17] | 16<br>(13.3) | 1.39<br>[0.78, 2.48] |
| g. It is better to become immune to a disease by getting sick than by getting a shot | 49<br>(1.7) | 11<br>(8.1) | 4.26<br>[2.14, 8.47] | 40<br>(4.3) | 2.87<br>[1.85, 4.44] | 29<br>(6.4) | 3.73<br>[2.28, 6.11] | 12<br>(4.7) | 2.82<br>[1.42, 5.57] | 10<br>(8.3) | 5.45<br>[2.66, 11.16] |
| h. I worry that government rushed the vaccine approval process | 577<br>(20.1) | 50<br>(36.5) | 2.10<br>[1.44, 3.05] | 311<br>(33.7) | 1.81<br>[1.53, 2.15] | 159<br>(34.8) | 1.68<br>[1.34, 2.10] | 76<br>(29.6) | 1.28<br>[0.96, 1.72] | 35<br>(29.7) | 1.31<br>[0.85, 2.00] |
| **Worries about vaccine (worries), % strongly agreeing or agreeing** |  |  |  |  |  |  |  |  |  |  |  |
| Might not stop you from getting COVID | 814 | 53 | 1.55 | 361 | 1.60 | 189 | 1.73 | 88 | 1.24 | 47 | 1.40 |
|  | (28.0) | (38.1) | [1.08, 2.22] | (38.4) | [1.37, 1.88] | (40.9) | [1.40, 2.13] | (34.1) | [0.94, 1.63] | (38.2) | [0.95, 2.05] |
| Might give you COVID and make you sick | 367<br>(12.7) | 48<br>(35.3) | 3.22<br>[2.20, 4.70] | 236<br>(25.2) | 2.45<br>[2.02, 2.96] | 138<br>(29.9) | 2.48<br>[1.95, 3.15] | 56<br>(21.8) | 1.73<br>[1.24, 2.40] | 41<br>(33.9) | 3.37<br>[2.25, 5.05] |

aOR=adjusted Odds Ratio, from logistic regression models including covariates for age, gender, cohort, and education. Reference group is white.

**Table 4:**
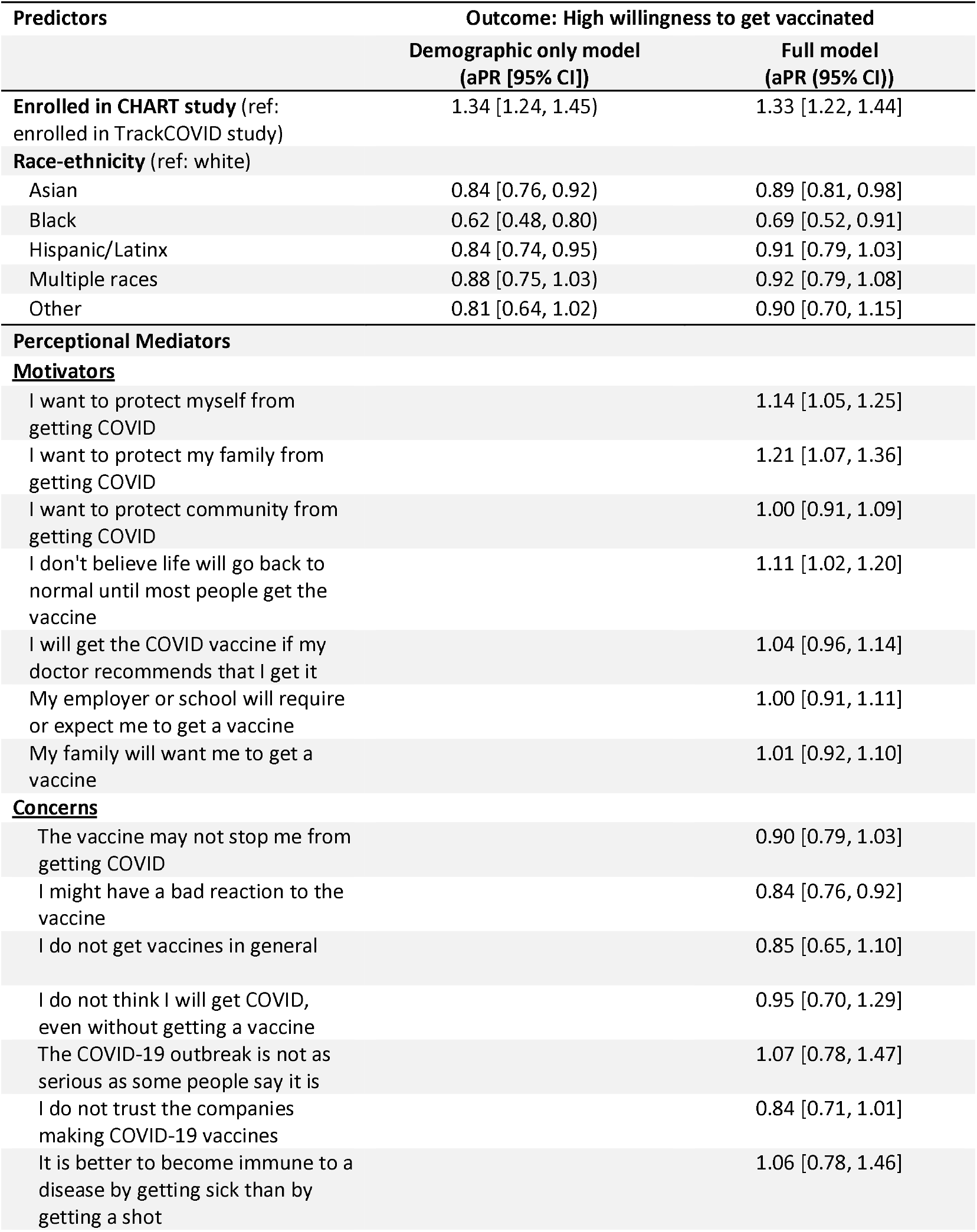
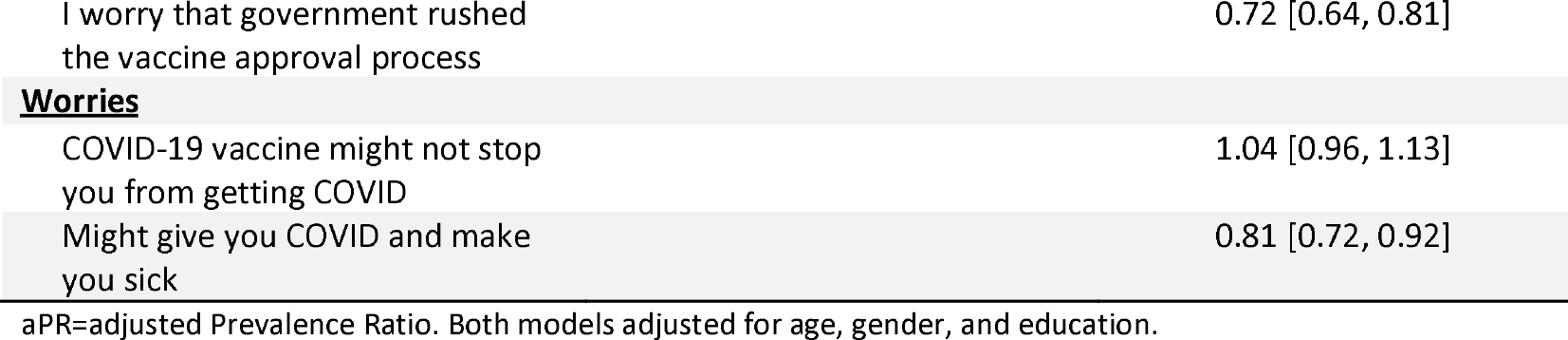
Perceptional factors (motivators, concerns and worries) mediators for race-ethnicity disparity in Vaccine Willingness.

| Predictors | Outcome: High willingness to get vaccinated |  |
| --- | --- | --- |
|  | Demographic only model<br>(aPR [95% CI]) | Full model<br>(aPR (95% CI)) |
| **Enrolled in CHART study** (ref: enrolled in TrackCOVID study) | 1.34 [1.24, 1.45] | 1.33 [1.22, 1.44] |
| **Race-ethnicity** (ref: white) |  |  |
| Asian | 0.84 [0.76, 0.92] | 0.89 [0.81, 0.98] |
| Black | 0.62 [0.48, 0.80] | 0.69 [0.52, 0.91] |
| Hispanic/Latinx | 0.84 [0.74, 0.95] | 0.91 [0.79, 1.03] |
| Multiple races | 0.88 [0.75, 1.03] | 0.92 [0.79, 1.08] |
| Other | 0.81 [0.64, 1.02] | 0.90 [0.70, 1.15] |
| **Perceptual Mediators** |  |  |
| **<u>Motivators</u>** |  |  |
| I want to protect myself from getting COVID |  | 1.14 [1.05, 1.25] |
| I want to protect my family from getting COVID |  | 1.21 [1.07, 1.36] |
| I want to protect community from getting COVID |  | 1.00 [0.91, 1.09] |
| I don't believe life will go back to normal until most people get the vaccine |  | 1.11 [1.02, 1.20] |
| I will get the COVID vaccine if my doctor recommends that I get it |  | 1.04 [0.96, 1.14] |
| My employer or school will require or expect me to get a vaccine |  | 1.00 [0.91, 1.11] |
| My family will want me to get a vaccine |  | 1.01 [0.92, 1.10] |
| **<u>Concerns</u>** |  |  |
| The vaccine may not stop me from getting COVID |  | 0.90 [0.79, 1.03] |
| I might have a bad reaction to the vaccine |  | 0.84 [0.76, 0.92] |
| I do not get vaccines in general |  | 0.85 [0.65, 1.10] |
| I do not think I will get COVID, even without getting a vaccine |  | 0.95 [0.70, 1.29] |
| The COVID-19 outbreak is not as serious as some people say it is |  | 1.07 [0.78, 1.47] |
| I do not trust the companies making COVID-19 vaccines |  | 0.84 [0.71, 1.01] |
| It is better to become immune to a disease by getting sick than by getting a shot |  | 1.06 [0.78, 1.46] |

|  |  |
| --- | --- |
| I worry that government rushed the vaccine approval process | 0.72 [0.64, 0.81] |
| **<u>Worries</u>** |  |
| COVID-19 vaccine might not stop you from getting COVID | 1.04 [0.96, 1.13] |
| Might give you COVID and make you sick | 0.81 [0.72, 0.92] |

aPR=adjusted Prevalence Ratio. Both models adjusted for age, gender, and education.

Black, Latinx, and Asian respondents were more likely than white respondents to endorse reasons to not get vaccinated (Table 3). Compared with whites, about 15% more of the Black, Latinx, and Asian respondents endorsed “bad reaction to the vaccine” and “government rushing the approval process” as major reasons to not get vaccinated. About one quarter of Black respondents endorsed lack of trust in the companies making COVID-19 vaccines and in the vaccine preventing COVID-19 as major reasons not to get vaccinated. Black, Latinx, Asian, multi-racial, and other race respondents were much more likely than white respondents to report concerns about the vaccine giving them COVID-19.

When comparing motivators, concerns and worries according to study cohort rather than race-ethnicity, CHART and TrackCOVID respondents had comparable rankings of the most important reasons to get, or not get, vaccinated (Appendix Table 1).

### Perceptional Factors (Motivators, Concerns and Worries) as Mediators for the Association between Race-ethnicity and Vaccine Willingness

Perceptional factors with significant positive association with high willingness to be vaccinated were a desire to protect family and one’s self from COVID-19 and belief that life will not get back to normal until most people get the vaccine (Table 4). The factors significantly and negatively associated with high willingness to get vaccinated were concerns about government rushing the approval process, getting a bad reaction from the vaccine, and getting COVID-19 from the vaccine. Comparing the adjusted prevalence ratios (aPR) for racial-ethnic groups for the first and second regression models indicates whether the motivators and concerns explain a portion of the association between cohort or race-ethnicity and high willingness to be vaccinated. The aPRs for each of the racial-ethnic groups are closer to 1 in the second model, suggesting that the perceptional factors included in the survey explain at least a small portion of lower vaccine willingness for Black, Latnix and Asian respondents relative to whites. Interestingly, the aPR for CHART Study participants remained virtually unchanged in the second model, suggesting a persistent association of being a CHART Study participant with vaccine intention that is not explained by differences between medical center employees and the general public in the motivations and concerns measured in the survey.

### Top Sociodemographic and Perceptional Predictors of Vaccination Willingness

The LASSO model identified the following as the most important factors in explaining COVID-19 vaccination intention: being in the CHART cohort, concern about government rushing the vaccine process, concern about having a bad reaction to the vaccine, motivation to protecting one’s self and family from COVID-19, and concern about getting COVID from the vaccine (Figure 1). The AUC of the LASSO model was 0.784 (95% CI: 0.783-0.822).

**Figure 1:**
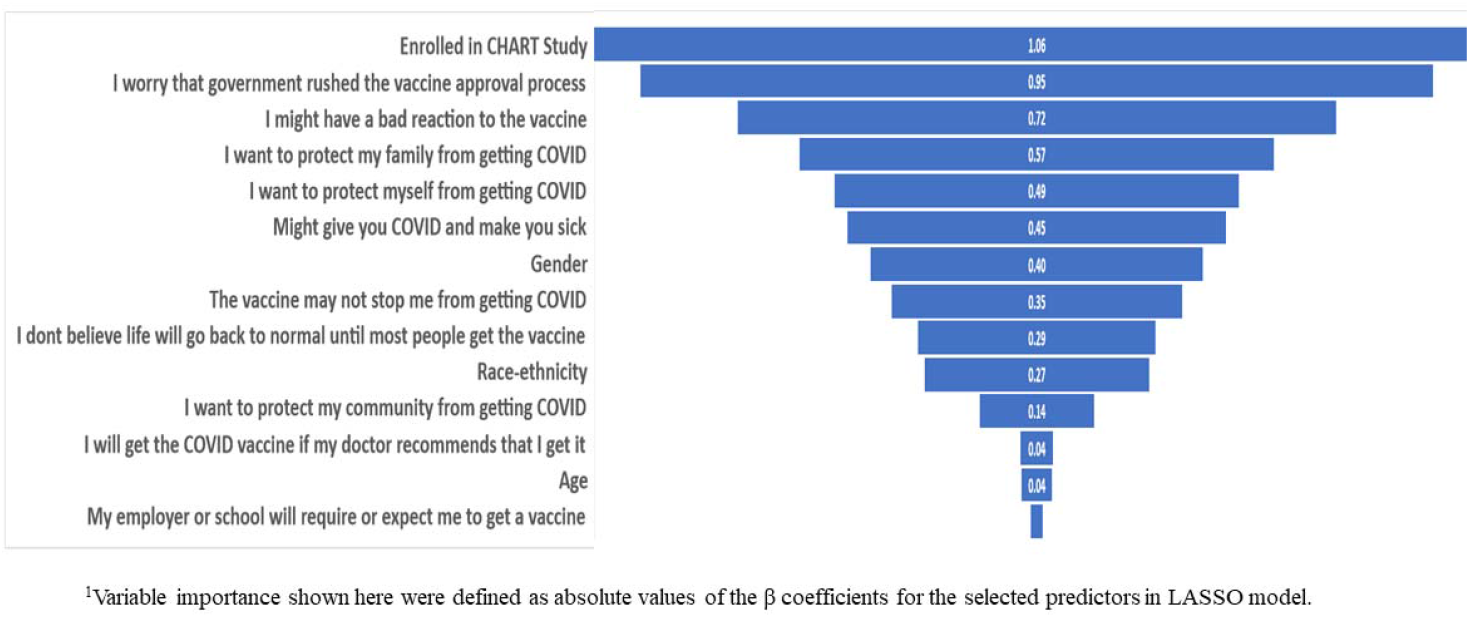
Relative Importance of Top Predictors for Vaccine Willingness, based on LASSO Regression Results^1^.

### Sensitivity Analyses

Patterns of associations between race-ethnicity, perceptions, and vaccination willingness were essentially the same when we used the alternative measure of high willingness, and the measure of low intention, as our outcome variables. For example, although the alternative measure of high willingness gave a lower point estimate of willingness than the measure included in the results reported above, both measures had similar associations with cohort, race-ethnicity, and vaccination reasons. Top predictors selected by LASSO were slightly different for the alternative measure of high willingness. The most important factors included (in rank order): concern about government rushing the vaccine process, concern about having a bad reaction to the vaccine, concern about getting COVID from the vaccine, race-ethnicity, and being in the CHART cohort (Appendix Figure 1).

## Discussion

Our findings are consistent with previous research that has found lower intention to receive a COVID-19 vaccine among Black and Latinx populations^2-9,18, 20-27^. Many of those studies were conducted early in the vaccine development stage, months before Emergency Use Authorization in the US of the first COVID-19 vaccines. Our survey period straddled the dates of emergency use authorization of the Moderna and Pfizer-BioNTech vaccines—a time when decision-making about vaccination was close to the start of actual vaccine availability for health workers but not for the majority of the US public. Our study sample is noteworthy for its breadth of racial-ethnic diversity, allowing us to report data separately for multiple racial-ethnic groups. We found that Asian, multi-racial, and other race-ethnicity respondents were more similar to Black and Latinx than white respondents in their intentions and beliefs about COVID-19 vaccination. Ours is one of the first published studies in the US to compare intentions among health workers and the general public in the same region directly, demonstrating that immersion in a health care setting did not offset racial-ethnic differences in intention.

Motivations for vaccination were comparable among respondents across all racial-ethnic groups in our study. Similar to findings from other studies, the most important reasons to get vaccinated were a desire not only to protect one’s self, but also one’s family and community^28^. The most prevalent concerns about COVID-19 vaccination among respondents to our survey were also similar to ones reported in other studies^20, 23-25, 27^, with concerns about efficacy and safety being most commonly endorsed. Whereas responses to reasons to get vaccinated were comparable across racial-ethnic groups in our study, Black, Latinx, and Asian respondents were much more likely than white respondents to endorse reasons to not get vaccinated, and in particular to endorse lack of trust as a reason to not get vaccinated.

Differences in motivations and concerns about COVID-19 vaccination across racial-ethnic groups only partly explained the observed racial-ethnic disparities in vaccine intentions, suggesting that there are additional unmeasured motivations and concerns across groups that mediate differences in vaccination intentions. Among the items measured, concern about a rushed vaccine approval process emerged as the single strongest predictor of intentions in both the regression and LASSO models, with a high concern being associated with lower willingness to get vaccinated. Trust in government has been found to be a factor associated with COVID-19 vaccine intentions among residents of many nations.^27-28^ Ours is one of the few published studies^3, 19, 24, 27^ to test the independent contribution of a broad range of beliefs as predictors of COVID-19 vaccination intentions. In addition, we applied both multivariate regression techniques and statistical learning methods to identify potential mediators of racial-ethnic differences as well as top predictors in COVID-19 vaccination intention.

Study limitations include sampling dates that were relatively early after the arrival of the Pfizer and Moderna vaccines to the US landscape. As such, they represent early views of healthcare workers and the general public and may not account for shifts in viewpoints, increasing confidence, or other changes in opinions in recent months as the US vaccination campaign has expanded greatly. Our survey sample was drawn from people sufficiently concerned about their risk of COVID-19 and trusting of research to volunteer for studies involving repeated COVID-19 testing. Self-selection may bias our results towards greater willingness to be vaccinated compared with the vaccine willingness of all medical center employees and community residents in the region studied. However, self-selection is less likely to introduce bias when testing associations of variables within the study cohorts, such as associations between race-ethnicity and vaccine intentions or between perceptional factors and intentions. It is striking that even within groups of individuals motivated to participate in longitudinal COVID-19 surveillance, large racial-ethnic differences exist in COVID-19 vaccination intentions and reasons not to get vaccinated. Our sample is from a single region in California, which may limit generalizability. Our survey may not have addressed certain domains that might influence intentions, such as concerns about access to vaccines. Our primary outcome was self-reported vaccine intention and not actual receipt of a vaccine. We plan to continue to study these cohorts to measure actual vaccination.

Our study has important implications for COVID-19 vaccination strategies. Our findings highlight that even for workers in the health sector, special effort is required to reach historically marginalized populations to support informed decision-making about vaccination. These campaigns must forthrightly acknowledge the history of racism in biomedical research and healthcare delivery that has injured race-ethnicity minority groups, degraded the trustworthiness of health and medical science institutions, and continues to undermine confidence in vaccines.^30,31^ Although our study suggests that educational outreach must address common concerns about vaccine risk and efficacy, it also points to positive messages that may resonate. Altruism was a strong motivator for vaccination, especially the desire to protect one’s family. Vaccination decision-making is a deliberative and dynamic process. This may be particularly salient for interpreting the powerful effect on intentions of concern about a rushed vaccine approval process. This concern may be mitigated over time as more people are vaccinated and evidence accumulates about vaccine efficacy and safety. Finally, it is important to emphasize that addressing motivations and concerns must not distract from the importance of ensuring more equitable access to vaccination^8,32^. Many individuals in all racial-ethnic groups have high willingness to be vaccinated but face barriers to obtaining vaccination^7,33^.

In summary, vaccine outreach campaigns must ensure that the disproportionate toll of COVID-19 on communities of color is not compounded by inequities in vaccination. Efforts must emphasize messages that speak to the motivations and concerns of groups suffering most from health inequities and ensure equitable access to vaccinations.

## Data Availability

Analytical data will be available with the approval of the California Pandemic Consortium upon request.

## Acknowledgments

California Pandemic Consortium: Lloyd Minor, MD; Anna Graber; Christopher Leung; Anthony Bet ; Matthew Sklar; Cole Holderman ; Amanda Kempema; Hilary Tang; Joseph Lohmann; Cole Holderman; Dan Lowenstein, MD; David Glidden; Rodolfo Villa; Charles Craik; Jessica Chao; Aida Julien; Marcus Paoletti; Sravya Jaladanki; Guntas Padda; Daisy Valdivieso; Lillian Brown; Carina Marquez; Jacob Ghahremani; Emerald Wan; Audrey Mustoe; Steve Miller; Ralph Gonzalez; Joe DeRisi; Sabrina Mann; Emily Crawford; CLIA HUB consortium; Parul Bhargava

The authors thank all project staff for their dedicated work on this study, the altruism of participants in both TrackCOVID and CHART study, and community partners who collaborated on recruitment of participants for the Track COVID cohort.

## Declarations

### Availability of data/material/code

All study materials are available for request upon approval of the corresponding author.

### Author Contributions

All authors contributed to the study conception and design. Material preparation, data collection and analysis were performed by Yingjie Weng, Di Lu and Kevin Grumbach. The first draft of the manuscript was written by Yingjie Weng and all authors commented on previous versions of the manuscript. All authors read and approved the final manuscript.

### Consent to participate

Informed consent were obtained from all study particiants.

---

**Apendix Table 1.**
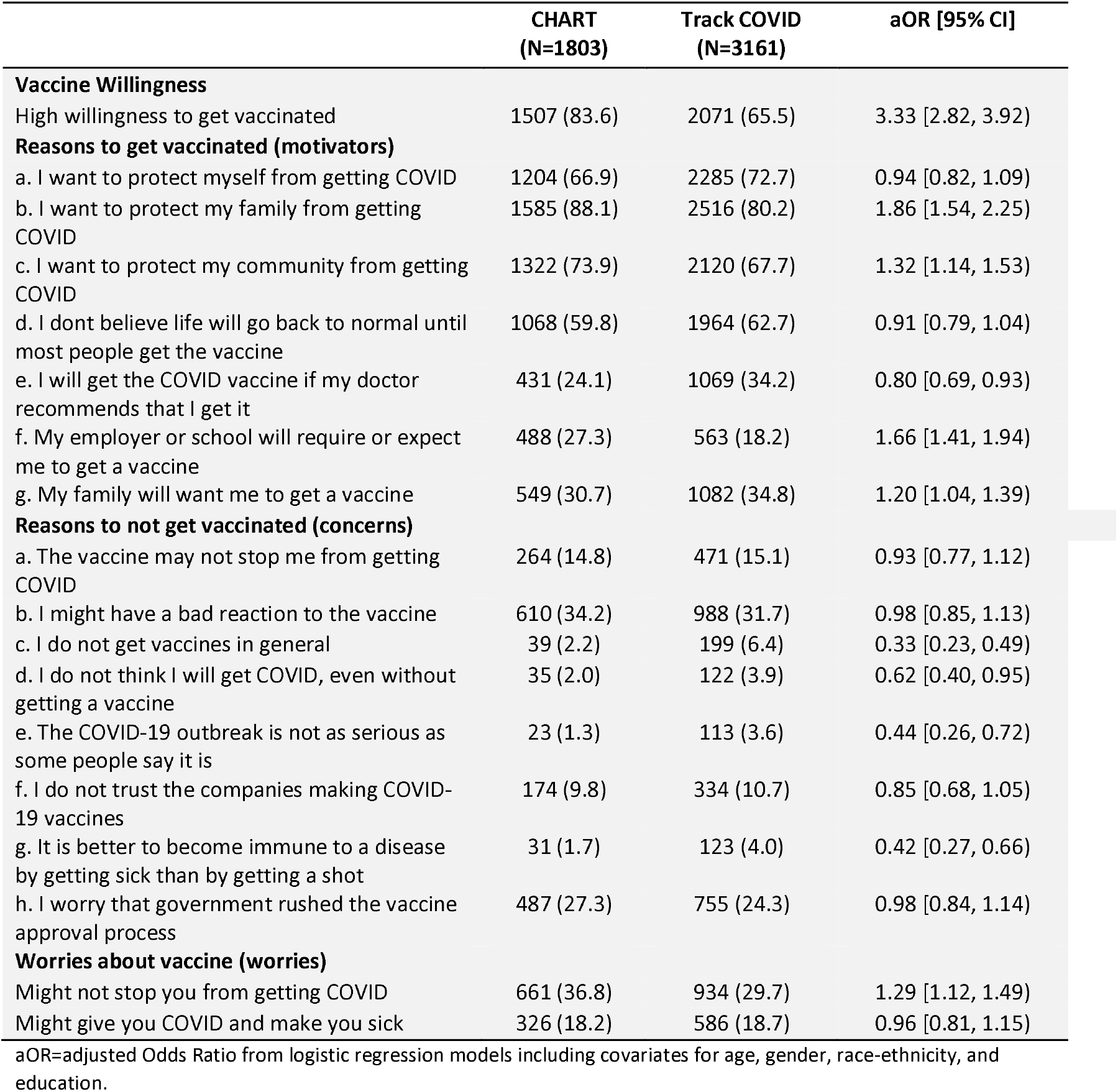
Vaccine willingness, motivators, and concerns by study cohorts (reference: Track COVID cohort)

**Apendix Table 2.**
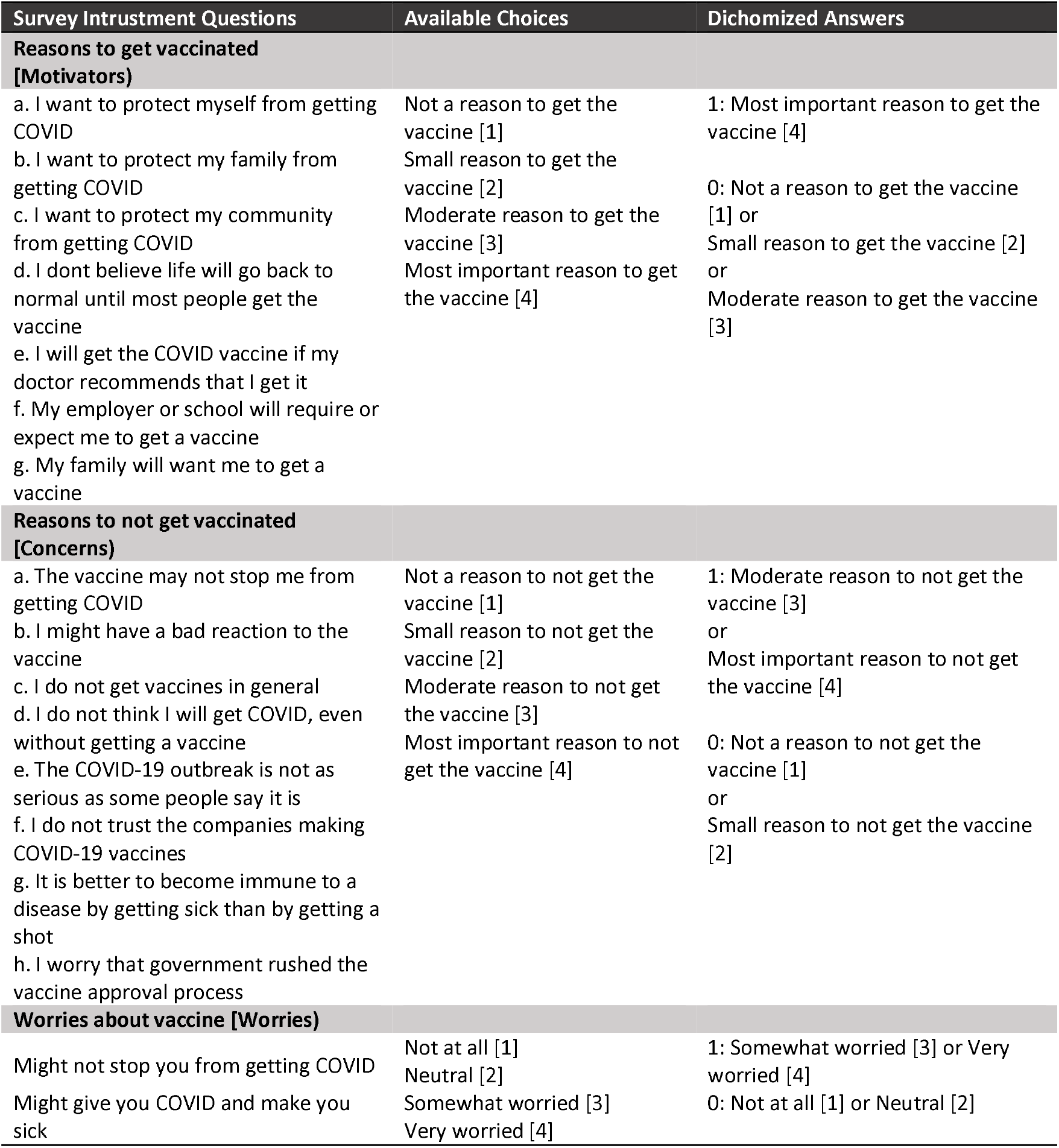
Survey Instruments on Perceptional Factors of COVID-19 Vaccine Willingness: Motivators, Concerns and Worries.

**Apendix Table 3.**
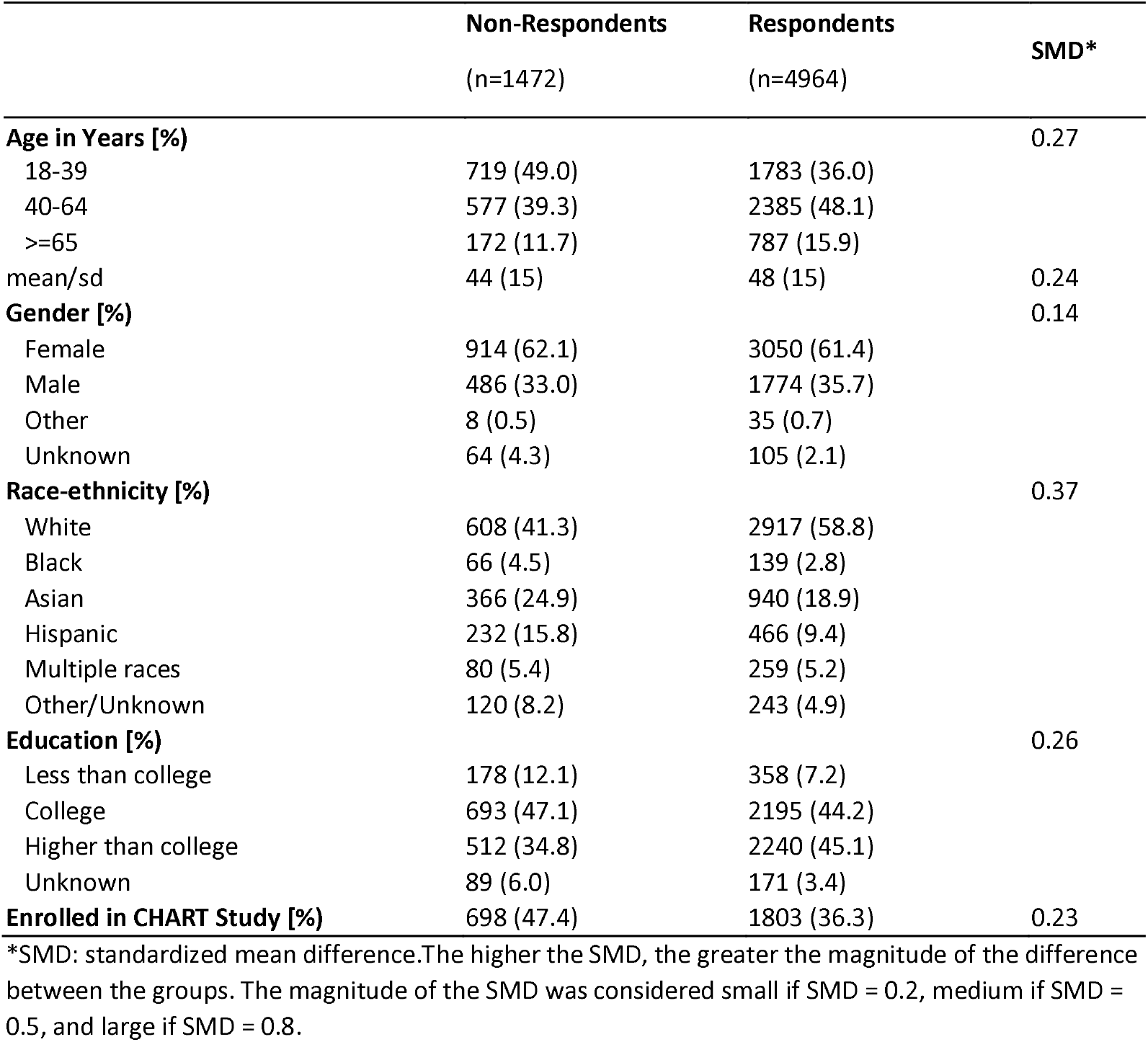
Demographic Characteristics of Vaccine Survey Respodents vs Non-respondents.

**Apendix Figure 1.**
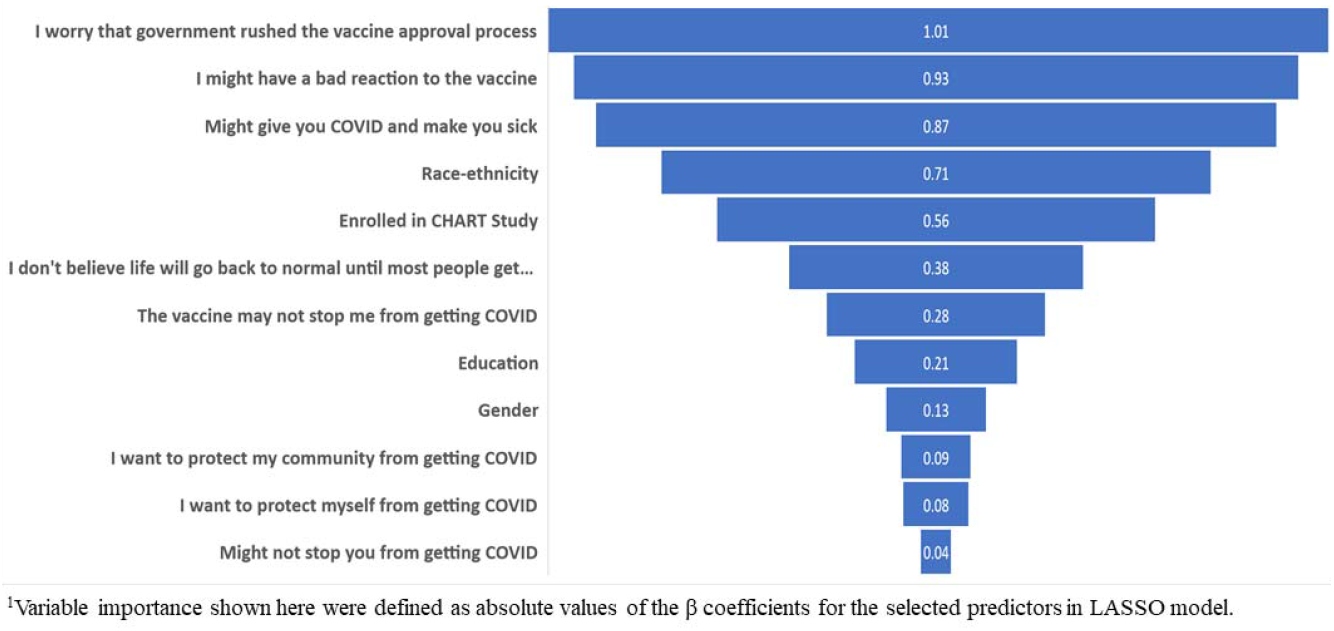
Variable Importance of Top Predictors for Vaccine Willingness [Altenative Measure] ^1^.

